# AI/ML Models to Aid in the Diagnosis of COVID-19 Illness from Forced Cough Vocalizations: Results and Challenges of a Systematic Review of the Relevant Literature

**DOI:** 10.1101/2021.11.12.21266271

**Authors:** K. Kelley, A.A. Sakara, M. Kelley, S. C. Kelley, P. McLenaghan, R. Aldir, M. Cox, N. Donaldson, A. Stogsdill, S. Kotchou, G. Sula, M.A. Ramirez

**Affiliations:** Chief Regulatory Medical Director for RAIsonance, Inc.; Co-Chief for Clinical and Regulatory Affairs for RAIsonance, Inc.; SVP Clinical Trials and Quality Improvement for RAIsonance, Inc.; SVP Market and Business Development for RAIsonance, Inc.; Chairperson Healthcare Provider Advisory Board for RAIsonance, Inc.; Lead AI/ML Biotechnical Engineer for RAIsonance, Inc.; AI/ML Biotechnical Engineer for RAIsonance, Inc.; Intern at RAIsonance, Inc.; Chief Medical Officer, Chief Scientist, Chief Strategy and Planning Officer for RAIsonance, Inc

## Abstract

From a comprehensive and systematic search of the relevant literature on signal data signature (SDS)-based artificial intelligence/machine learning (AI/ML) systems designed to aid in the diagnosis of COVID-19 illness, we aimed to reproduce the reported systems and to derive a performance goal for comparison to our own medical device with the same intended use. These objectives were in line with a pathway to regulatory approval of such devices, as well as to acceptance of this unfamiliar technology by disaster/pandemic decision makers and clinicians. To our surprise, none of the peer-reviewed articles or pre-print server records contained details sufficient to meet the planned objectives. Information amassed from the full review of more than 60 publications, however, did underscore discrete impediments to bringing AI/ML diagnostic solutions to the bedside during a pandemic. These challenges then were explored by the authors via a gap analysis and specific remedies were proposed for bringing AI/ML technologies in closer alignment with the needs of a Total Product Life Cycle (TPLC) regulatory approach.

## 1. Introduction

The onset of the SARS-CoV-2 outbreak in Wuhan, China in December 2019 and subsequent WHO declaration of a global pandemic in March 2020 has rallied workforces of every skillset to the cause. Frontline healthcare teams, supply chain employees, educational staff, and utilities personnel were called upon as Essential Workers. Anticipating the needs for processing large amounts of pandemic-related data, the Artificial Intelligence/Machine Learning (AI/ML) and Data Science (DS) communities also joined forces to contribute their talents to the response effort. AI/ML technologies were applied to the development and maintenance of numerous types of signal data signature (SDS) libraries, registries, and clinical datasets from forced cough vocalizations (FCV). Voice, breath, and FCV research already had established that neural networks coupled with feature extraction analysis by AI/ML engines could be used to identify a variety of respiratory and neurological diseases. Drawing from this prior body of work, teams across the globe began working independently to establish SDS-based software systems to aid in the diagnosis of COVID-19 illness. But how many of the resulting software-as-medical-devices (SaMDs) would have the safety and performance profiles to support a viable regulatory pathway to market? And would the application and usability of this unfamiliar technology gain acceptance for clinical use during a disaster response?

To answer these questions, we prospectively planned a PRISMA 2020 systematic review of the relevant literature. Our primary objectives were to demonstrate reproducibility of the published models by building each COVID-19 diagnostic software system for which sufficient details were reported and to conduct head-to-head evaluations both across the completed models and also in comparison to our own AI/ML system. As a secondary objective, our intent was to determine a literature-derived performance goal (PG) for the completed models for comparison to our device with the same intended use.

## 2. Background

Implementation of AI/ML solutions to clinical needs hinges upon strong adherence to scientific principles, good clinical practice, and the medical device regulatory process. Specific to times of public health emergency, numerous governments and regulatory agencies have authority to bring devices, drugs or biologics with a risk-to-benefit profile deemed acceptable into use for affected populations. The US-FDA Emergency Use Authorization (EUA), the WHO Emergency Use Letter (EUL), and a global array of expedited governmental and healthcare agency review pathways are all examples of routes to emergency clinical use. And key to achieving rapid acceptance of and fostering trust in newer technologies during disasters or emergencies is keeping the regulatory process central to all aspects of design, development, and deployment. In other words, documentation spanning the Total Product Life Cycle (TPLC) from inspiration to post-market surveillance.

In response to the immense need for COVID-19 diagnostic testing during the pandemic, scientific teams of diverse disciplines have developed hundreds of devices, most of which were based upon established methods of laboratory analysis. There were other approaches, though, proposed by AI/ML and Data Science professionals in search of a scalable diagnostic device that did not require wet specimens or laboratory processing. Academic institutions from around the world have cataloged, analyzed, and published peer-reviewed articles on SDS libraries that were amassed both before and after December 2019. This collective wealth of data contains FCV recordings from years prior to the pandemic (not COVID-19), post-December 2019 recordings of COVID-19 negative persons as confirmed by RT-PCR (not COVID-19), and FCV recordings from persons diagnosed COVID-19 positive and verified by RT-PCR (yes COVID-19). While some author groups employed in-house collections of recordings, there are a number of publicly available SDS libraries that are identifiable from literature searches and downloadable for use in the development of SDS-based COVID-19 diagnostic software systems.

## 3. Materials and Methods

Certain terms will be adopted uniformly throughout this review, to provide clarity. The term “article” will be reserved for peer-reviewed references and those obtained from pre-print servers (non-peer-reviewed) will be termed “records”. While many of the references refer to an SDS collection from FCV as a “registry”, the reviewers will use the term “library”, so as not to suggest that the same degree and quality of clinical data typically included in medical device registry exists in each SDS collection. Lastly, the shorter phrase “SDS Library” will be representative of each SDS library from FCV.

### 3.1. Literature Searches

On 12 October 2021 and updated on 7 November 2021, we conducted systematic searches of the relevant literature for the purpose of presenting a comparative evaluation of AI/ML systems designed to aid in the diagnosis of COVID-19 from FCV. Searches of the peer-reviewed literature were prioritized but, given the collaborative “shareware” culture of the AI/ML and Data Science communities, pre-print servers were searched for possible contributions. EndNote 2020 was the designated reference manager and PubMed was searched via this software. Serial searches of “Any Field” in PubMed, “Full Text and Metadata” in the IEEE Xplore digital library of the Institute of Electrical and Electronics Engineers,(ieeexplore.ieee.org), “All Fields” in the arXiv open-access archive (arxiv.org), and “Full Text or Abstract or Title” in bioRxiv and medRxiv (medrxiv.org) were performed using the identical search terms as listed below:

- covid and classifier
- covid and neural network
- covid and cough and artificial intelligence
- covid and cough and AI
- covid and cough and machine learning
- covid and cough and ML
- covid and cough and classifier
- cough and neural network
- forced cough vocalization

The results from these serial searches were combined and systematically filtered to achieve a final article pool from which all references would be evaluated for contribution to the stated objectives. Following a basic PRISMA 2020 workflow, our search methodology is illustrated below in Figure 1:

**Figure 1.**
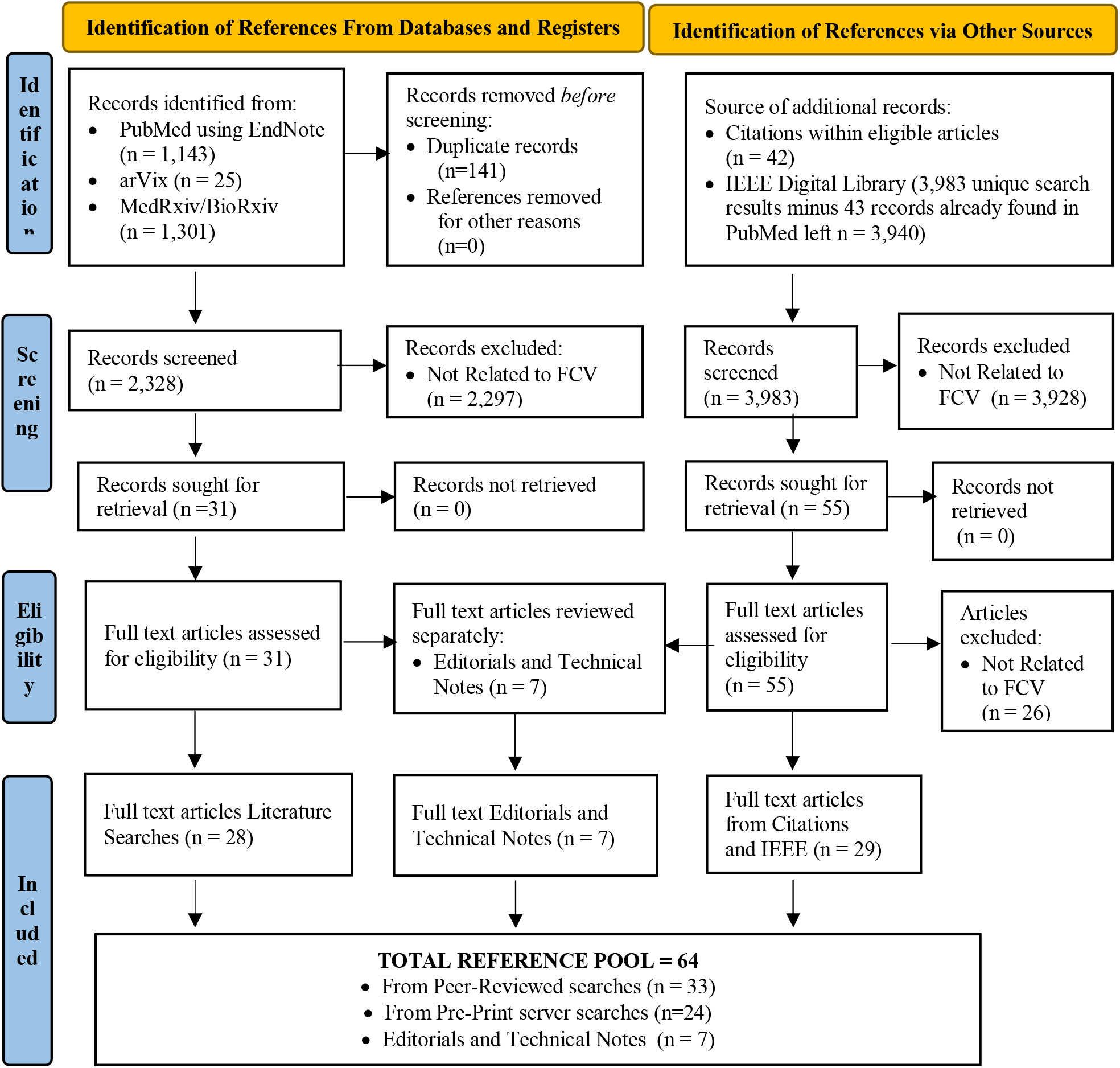
PRISMA 2020 Flow Diagram for New Systematic Reviews (inclusive of Databases, SDS Libraries, and Other Sources)

### 3.2. Classification System for SDS Libraries

Since the introduction of levels of evidence into the scientific literature, many professional organizations and journals have adopted some variation of this classification system. Diverse specialties are often asking different questions, though, and it has been recognized that the type and level of evidence needs to be modified accordingly [1]. Evidence-Based Medicine classification systems specifically are based upon research design questions that take into account prospective vs retrospective data collection, data collection methods, data sources, data verification, control design, study population sampling, diagnosis, and decision analysis. In order for comparisons to be made across the many SDS libraries used in the above references, an appropriate classification system was required.

Grades of Data are another classification system, derived from the Level of Evidence (data collection design) and modified in concert with the same research design questions, but only as they apply to each data subset. Therefore, an SDS library with a Level 2b design is expected to yield Moderate Grade data. If some retrospective data were to be included in a given library, the inclusion of this data subset would reduce the grade level for the overall collection by one grade level to Low Grade data. Likewise, if some or all of the retrospective data also had incomplete clinical data, then the grade of data would be reduced by two levels (one each for retrospective and incomplete clinical data), degrading the library data to Insufficient [2, 3]. Conversely, if an SDS library with a Level 2b design yielding Moderate Grade data were to have a subset verified by two RT-PCR tests within 24 hours of the FCV recording, that subset of data would be classified as High Grade Data and the entire library would be elevated by one data grade.

To permit assignment of design levels and stratification of data as reported, the Levels of Evidence design and associated Grades of Data criteria were modified according to the needs of AI/ML training, validation, and testing sets. This yielded a classification system appropriate to the topic of our review and is shown below as Table 1.

**Table 1.** Levels of Evidence and Grade of Data Classification System for SDS Libraries.

| Design Level | Data Grade | Library Design | SDS Collection | Lab Confirmation | Clinical Confirmation | Possibly Elevated by Mined Data? * |
| --- | --- | --- | --- | --- | --- | --- |
| 1a | High | Prospective | Systematic | 2 tests within 24 hours of SDS | Yes | No |
| 1b | High | Prospective | Systematic |  | No | No |
| 1c | High | Prospective | Systematic | 1 test within 24 hours of SDS | No | Yes |
| 2a | Moderate | Prospective | Systematic |  | Yes | No |
| 2b | Moderate | Prospective | Systematic |  | No | No |
| 2c | Moderate | Prospective | Crowd-Sourced |  | No | Yes |
| 3a | Low | Prospective | Crowd-Sourced | 1 test > 24 hours of SDS | Yes | No |
| 3b | Low | Prospective | Crowd-Sourced |  | No | No |
| 4 | Insufficient | Retrospective | Wiki-Sourced | No | No | No |
| 5 | Insufficient | Retrospective | Synthetic |  | No | No |

\* higher grade data mined from lower design levels may be incorporated to elevate the overall data grade

## 4. Results

From the final pool of references, all were read and evaluated across a multidisciplinary team of Data Scientists, Biotechnical Engineers, Healthcare Clinicians, Product Development and Clinical/Regulatory Affairs professionals. There were 57 references proposing a specific COVID-19 diagnostic AI/ML model or application (33 peer-reviewed articles and 24 pre-print server records) and the remaining 7 articles were Editorials and Technical notes reserved for potential contribution to the Discussion.

### 4.1. Primary Objectives

Although each of the 57 references purported to include a fully-detailed AI/ML solution, only 14 contained enough information for us to attempt building the stated solution. Unfortunately, no single reference included sufficient details to build a complete model or system, rendering comparisons across models (including our own) unattainable. The most common omissions pertained to the model’s architecture or the flow of data through the model’s layers. Thus, the primary objectives of this review could not be met.

### 4.2. Secondary Objective

Determination of a literature-derived PG was the secondary objective of this review and is a familiar endpoint in regulatory pathways. To realize this goal, the intent was to mine the data as reported and as trained for the individual models and calculate the PG. AI/ML software solutions meeting the 5 prospective criteria as below, defining a comparable population, were to be included in the calculation:

- Commonly defined sensitivity and specificity of the model when tested with the respective test set
- False positive and false negative were reported in some fashion
- Test sets were of statistically significant size, based upon Exact Binomial Test
- Data sets represented good quality data, defined as a Design Level 1 or 2 and Data Grade of High or Moderate
- Results for PPA and NPA (directly reported or calculable from the details presented in the reference) were compared to RT-PCR for COVID-19 test results, in keeping with submission criteria for US-FDA EUA or WHO EUL

There were 6 of 57 references found to have a Level of Evidence design of 2 or better, High or Moderate Grade data, and a statistically significant SDS library size provided the testing results to meet criteria (see Table 2). And of these same 6 references, only 2 were peer-reviewed articles. None of the reference met all criteria, rendering the secondary objective unattainable. Additionally, no more than 2 of the 6 references shared a common endpoint, disabling even that aspect of potential comparison.

**Table 2.** Articles Meeting 3 of 5 Criteria for Establishing a Literature-Derived Performance Goal.

| Author | FCV/SDS Library Name | Level of Evidence (Collection Design) | Grade of Data Collected |
| --- | --- | --- | --- |
| **FROM PEER-REVIEWED ARTICLES** |  |  |  |
| Andreu-Perez et al. [4] | In-House Collection | 2a | Moderate |
| Verde et al. [5] | Coswara | 2a | Moderate |
| **FROM PRE-PRINT SERVER RECORDS** |  |  |  |
| Alkhodari et al. [6] | Coswara | 2a | Moderate |
| Chang et al. [7] | Coswara DiCORA Cough Sub-Challenge | 2a | Moderate |
| Chetupali et al. [8] | Coswara | 2a | Moderate |
| Muguli et al. [9] | Coswara | 2a | Moderate |

### 4.3. Gap Analysis

We previously had developed a categorization system specific to our objectives and determined the level of evidence and grade of data for each of the 57 references. To emphasize aspects of each library population rather than individual articles, this time we reorganized the information as seen below in Table 3:

**Table 3.** SDS Libraries Represented in Current Literature Search Article Pool.

| FCV/SDS Library Name | Author | Level of Evidence (Collection Design) | Grade of Data Collected |
| --- | --- | --- | --- |
| **FROM PEER-REVIEWED ARTICLES** |  |  |  |
| Coswara | [10], [11], [12], [5] | 2a | Moderate |
| In-House Collection | [13] | 2a | Moderate |
| In-House Collection | [4] | 2a | Moderate |
| In-House Collection | [14] | 2a | Moderate |
| In-House Collection | [15] | 2a | Moderate |
| In-House Collection | [16] | 2a | Moderate |
| In-House Collection | [17] | 2a | Moderate |
| In-House Collection | [18-20] | 2a | Moderate |
| In-House Collection | [21] | 2a | Moderate |
| Israeli COVID-19 Dataset | [22] | 2a | Moderate |
| Cambridge COVID-19 Cough Database | [23], [24], [25], [12] | 3a | Low |
| Carnegie Melon COVID-19 Voice Detect | [26] | 3a | Low |
| Corona Voice Detect | [27] | 3a | Low |
| COUGHVID | [23], [28] | 3a | Low |
| ICBHI Dataset 2017 | [29] | 3b | Low |
| MIT Open Voice Dataset COVID-19 Cough | [23], [30] | 3a | Low |
| NYU COIVD Cough Dataset | [23] | 3a | Low |
| Pertussis dataset | [12] | 3a | Low |
| Environmental Sound Classification (ESC-50) | [31] | 3b | Low |
| FreeSound | [32, 33], [34] | 3b | Low |
| Novel Corona Virus Dataset 2019 (Kaggle) | [35] | 3b | Low |
| Sarcos Dataset | [11] | 3b | Low |
| Univ. of Lleida | [12] | 3b | Low |
| Vgg-16 | [36] | 3b | Low |
| Virufy | [37], [38, 39], [12] | 3b | Low |
| Google Audio Set | [34] | 4 | Insufficient |
| Instagram | [40] | 4 | Insufficient |
| NoCoCoDa | [33], [38] | 4 | Insufficient |
| Twitter | [40] | 4 | Insufficient |
| YouTube | [40] | 4 | Insufficient |
| DCASE | [32, 33] | 5 | Insufficient |

| FROM PRE-PRINT SERVER RECORDS |  |  |  |
| --- | --- | --- | --- |
| Coswara | [6], [41], [42], [43, 44], [45] | 2a | Moderate |
| Coswara DiCOVA Cough Sub-Challenge | [46], [7], [9] | 2a | Moderate |
| In-House Collection | [47] | 2a | Moderate |
| In-House Collection | [48] | 2a | Moderate |
| In-House Collection | [43] | 2a | Moderate |
| In-House Collection | [49] | 2a | Moderate |
| AICovidVN 115M | [50] | 3a | Low |
| Brooklyn | [42] | 3a | Low |
| Cambridge COVID-19 Cough Database | [8], [41, 51], [46]] | 3a | Low |
| ComPare CSS | [46], [42], [52, 53] | 3a | Low |
| COUGHVID | [46], [8] | 3a | Low |
| COVID-19 Sounds Dataset | [45] | 3a | Low |
| MIT Open Voice Dataset COVID-19 Cough | [47], [8], [45] | 3a | Low |
| TASK dataset | [42] | 3a | Low |
| Wallacedene dataset | [42] | 3a | Low |
| Environmental Sound Classification (ESC-50) | [54], [55], [56] | 3b | Low |
| Crowdsourced Respiratory Sound Data | [48] | 3b | Low |
| Flusense (negatives) | [47], [43] | 3b | Low |
| FreeSound Database (negatives) | [47], [42], [43] | 3b | Low |
| Google Audio Set | [42] | 3b | Low |
| IATos | [57] | 3b | Low |
| Medina Medical Group Russia | [58] | 3b | Low |
| Sarcos Dataset | [42] | 3b | Low |
| Virufy | [51] | 3b | Low |
| NoCoCoDa | [51] | 4 | Insufficient |
| YouTube | [59] | 4 | Insufficient |

Looking at the review data from this vantage point, we observed that all of the SDS libraries collected by author groups themselves for the purpose of their model development (identified as In-House Collections) had Level 2a design that yielded Moderate Grade data. And only the projects employing the Coswara library (whether or not the reference was peer-reviewed) and the Israeli COVID-19 dataset ranked equally.

Since the intended objectives could not be met, we reexamined the data from our review for alternative utility. The logical next steps from a regulatory approach were to complete a formal gap analysis, to focus on the issues that rendered our aims unattainable. Perhaps the challenges central to researching diagnostic AI/ML solutions for COVID-19 were representational of the greater challenges faced by AI/ML development teams entering the medical device market, particularly during a public health emergency? Answering this question and itemizing the contributing factors became our new objectives.

The results of our gap analysis are summarized below in Table 4, itemizing the recurring themes identified in the body of literature assembled during this review. Each gap directly impacted our ability to meet primary and secondary objectives but also would affect the market-readiness of any medical device. Plans proposed here to mitigate or resolve these gaps were intended to apply not only to future AI/ML development publications but also to support realization of a regulatory pathway, keeping the needs of the TPLC in mind and clinical relevance at the core.

**Table 4.** Gap Analysis Summary.

| **Current State** | **Ideal State** | **Gap** | **Remedy** |
| --- | --- | --- | --- |
| Low quality data within available COVID-19 SDS libraries/registries | High quality data | Lack of an evidence-based system of data categorization | Employ a system that combines familiar medical Levels of Evidence and Grading of Data into a single architecture for the evaluation of SDS data |
|  |  | Inherent subjectivity of symptom reporting and clinical diagnosis framing bias | Include objective measures of illness to optimize the Grade of Data |
| Undersized datasets for pre-trial training, validation, and testing | Adequate sizing of all datasets | Lack of statistical significance of datasets | Ensure that datasets are statistically significant prior to clinical trial design (key to calculating sufficient trial population size and trial endpoints) |
| None of the AI/ML model articles reported or proposed a plan for deployment in the clinical world | Incorporation of clinically relevant steps for deployment early in the design, development, and testing processes | Focus of reports are on software design and development, often without clear correlation to clinical application | Include input from a multidisciplinary team, including regulatory, across the spans of the TPLC |
| None of the models met the minimum bar for emergency use consideration by FDA or WHO | Models and pathways to regulatory submission are compared to current standards for safety and performance | Minimum criteria of regulatory standards not met, as reported in the references | Compare each stage of the TPLC to regulatory standards, demonstrating that all criteria were attained or are attainable |
| Models could not be built as reported | Adhere to the scientific process of data validation, verification, and reproducibility | Incomplete documentation of codesets, datasets, and code implementations | Report complete documentation or at least provide the resulting transfer learning, to enable cross-checking of outcomes |

**Table 4:** Summary of Andreu-Perez et al. [4].

| STRENGTHS | CHALLENGES |
| --- | --- |
| Technically robust AI/ML system | Outcome is not a definitive diagnosis (only a probability of likely or unlikely COVID-19) |
| Platform independent | Incompletely documented |
| Broadly accessible web-based format | Web-based systems cannot turn off hardware-based sound processing (alters the source SDS) |
| GDPR compliant | Design for anonymity did not include maintenance of patient records or mandatory reporting |

**Table 5:** Summary of Verde et al. [5].

| STRENGTHS | CHALLENGES |
| --- | --- |
| Expressed a desire for a mobile healthcare solution via a smartphone | Did not report how the SVM algorithm would be embedded into a mobile health solution |
| Emphasized importance of conducting a controlled clinical trial to validate the AI/ML system | Focus of study was the algorithms and the data, rather than a deployable holistic solution |

**Table 6:**
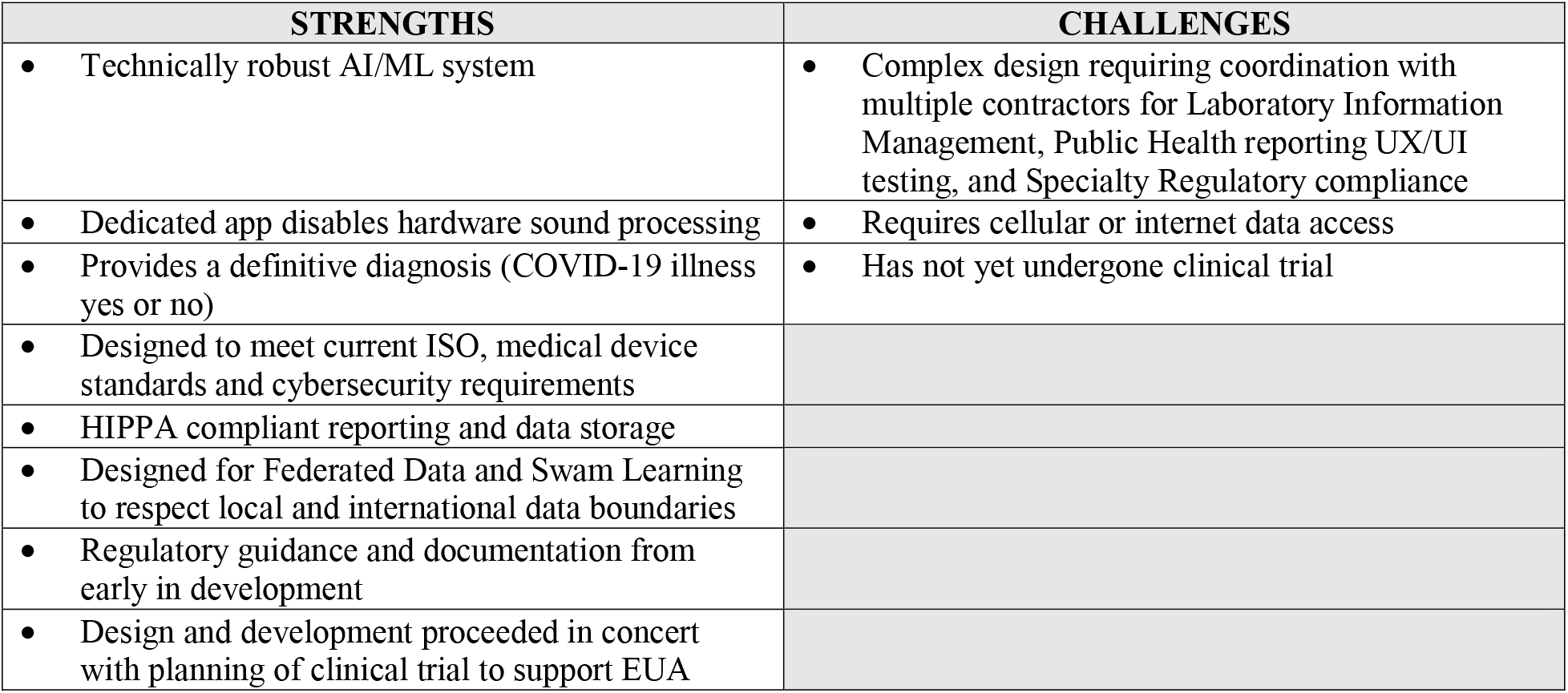
Summary of Our System in Development (Pre-Trial)

| STRENGTHS | CHALLENGES |
| --- | --- |
| Technically robust AI/ML system | Complex design requiring coordination with multiple contractors for Laboratory Information Management, Public Health reporting UX/UI testing, and Specialty Regulatory compliance |
| Dedicated app disables hardware sound processing | Requires cellular or internet data access |
| Provides a definitive diagnosis (COVID-19 illness yes or no) | Has not yet undergone clinical trial |
| Designed to meet current ISO, medical device standards and cybersecurity requirements |  |
| HIPPA compliant reporting and data storage |  |
| Designed for Federated Data and Swam Learning to respect local and international data boundaries |  |
| Regulatory guidance and documentation from early in development |  |
| Design and development proceeded in concert with planning of clinical trial to support EUA |  |

## 5. Discussion

### 5.1. Employing Remedies to Bridge Gaps

Data quality issues and the ultimate impact on model function underscored the most common challenges noted in both the research and editorial articles contributing to this review [60], [61]. To enhance the data quality and optimize the Grade of data, several authors suggested methods to verify symptomology and augment clinical diagnosis of subjects with suspected COVID-19 illness. Specific technologies cited for improving library and data collection design include Internet-Of-Things (IOT) sensors, mobile digital health products, and mHealth wearable devices [62]. [60] [63], [61], [64], [65], [66].

### 5.2. Review of Level 2a, Moderate Grade, Statistically Significant Articles

At first glance, the reference pool identified by our searches and reviews of the literature appeared to yield relatively complete descriptions of working COVID-19 diagnostic AI/ML models available for download or as a live working model via the Internet. Re-creation of these models began as a promising endeavor, despite the need for training, validation, and testing specifics. Each was built by our teams according to the plans presented in the respective reference but, regrettably, none of the models were reproducible as a fully deployed or fully deployable device.

The recurring theme throughout our review process was that most references, at least as documented, provided more exploration than execution of diagnostic SaMD models for COVID-19. Some notable exceptions were the technological contributions offered in articles by Imran et al. [31] and Orlandic et al [28] and also the pre-print paper by Chowdhury et al. [41]. But without inclusion of a clear regulatory pathway, the authors did not bring the TPLC full-circle for the models they presented [41, 51] [28, 31]. Two additional publications of distinction were Andreu-Perez et al. [4] and Verde et al. [5]. As the only articles meeting a majority of criteria for establishing a performance goal for the AI/ML systems of interest, their respective work is summarized below and accompanied by a table of model-specific strengths and challenges.

Andreu-Perez, et al [4] reported on an AI/ML system to screen for COVID-19 based upon FCV. The SDS library employed was an in-house library collected in Spain, which was categorized in the current review as Level 2a design and a statistically significant amount of Moderate Grade data. Each SDS record was analyzed by the ML and SDS processing system, filtered by a cough detector, and results are given to the user. Incoming recordings were filtered, cleaned, and then passed through their cough detector based on empirical mode decomposition (EMD). Cough information then was transformed into tensor form using Mel-Frequency Cepstral Coefficients (MFCCs), Mel-scaled spectrograms, and Linear Predictive Coding Spectrum (LPCS) coefficients. Extracted from each SDS were 33 features which were mapped onto a 3D tensor, using a CNN with three main layers stacked four times. The authors achieved a true positive accuracy of 97.18% and a true negative rate of 96.64%. However, since false positive and false negative data were not reported, Positive Percent Agreement (PPA) and Negative Percent Agreement (NPA) could not be determined.

In the second of two AI/ML systems for in-depth discussion, Verde et al. [5] also employed Coswara as their source of SDS data. The authors analyzed a variety of ML algorithms to detect COVID-19 through the phonetic vowel sounds of /a/, /e/, and /o/ that are inherent to FCV as the feature for analysis. In their listing of the model performances, the Support Vector Machine (SVM) performed the best overall: achieved percentages for Accuracy, F1-Score, Specificity, Precision, and Recall were 97.07, 82.35, 97.37, 73.68, 93.33 respectively and Area Under the Curve = 0.954. But as with Andreu-Perez et al. [4], since false positive and false negative data were not reported, PPA and NPA were not calculable.

### 5.3. Introduction of Our System in Development

Our AI/ML system for the diagnosis of COVID-19 illness, in common with the Level 2a is based upon the SDS from FCV. Having determined the Level 2a design and Moderate Grade SDS libraries from a systematic review of the literature, we selected Coswara as the source for statistically significant sized datasets. Incoming recordings are filtered, cleaned, and then passed through a cough detector. Once the SDS has been declared an FCV through audio analysis, it is then segmented into individual FCV segments. This is completed using hidden Markov modeling, which detects the onset and completion of each FCV within a file, standardizing the FCV for the classification models. FCV information is then transformed into tensor form using Mel-Frequency Cepstral Coefficients (MFCCs), Mel-scaled spectrograms, and Fast Formant Transformation (FFT) to yield SDS for feature analysis. Each SDS feature is analyzed separately, extracted, and mapped using an ensemble based upon successful neural network modeling techniques of 2D-CNN and RNN in the peer-reviewed literature [4] [31] [28]. Our process also employs ML algorithms to detect COVID-19 illness via the phonetic vowel sounds inherent to FCV as the feature for analysis in a manner similar to Verde et al. [5]. In anticipation of our upcoming clinical trial, we have achieved a Positive Percent Agreement (PPA) greater than 0.90 and Negative Percent Agreement (NPA) greater than 0.85 in benchtop testing.

## 6. Conclusion

While unable to meet the intended objectives of this systematic review of the AI/ML system literature on SDS from FCV aiding in the the diagnosis of COVID-19 illness, the authors did complete a gap analysis that identified the principal issues and significant challenges facing this growing field of study. The consequences of such gaps directly affected not only the quality of data available in the relevant literature but also prevented analyses to assess regulatory readiness of the devices and applications presented. Acceptance of unfamiliar technology by disaster or pandemic decision makers and clinicians would require bridging of these gaps through adherence to a clear and well-documented TPLC approach to a viable regulatory pathway.

## Data Availability

All data produces is available upon request from the authors

## References

[1] Burns, P.B., R.J. Rohrich, and K.C. Chung, The Levels of Evidence and Their Role in Evidence-Based Medicine. Plastic and reconstructive surgery, 2011. 128(1): p. 305–310.

[2] Granholm, A., W. Alhazzani, and M.H. Møller, Use of the GRADE approach in systematic reviews and guidelines. British journal of anaesthesia, 2019. 123(5): p. 554–559.

[3] Schünemann, H.J., et al., Grading quality of evidence and strength of recommendations for diagnostic tests and strategies. Bmj, 2008. 336(7653): p. 1106–1110.

[4] Andreu-Perez, J., et al., A generic deep learning based cough analysis system from clinically validated samples for point-of-need covid-19 test and severity levels. IEEE Transactions on Services Computing, 2021.

[5] Verde, L., et al., Exploring the Use of Artificial Intelligence Techniques to Detect the Presence of Coronavirus Covid-19 Through Speech and Voice Analysis. IEEE Access, 2021. 9: p. 65750–65757.

[6] Alkhodari, M. and A. Khandoker, Detection of COVID-19 in smartphone-based breathing recordings using CNN-BiLSTM: a pre-screening deep learning tool. medRxiv, 2021.

[7] Chang, J., S. Cui, and M. Feng, DiCOVA-Net: Diagnosing COVID-19 using Acoustics based on Deep Residual Network for the DiCOVA Challenge 2021. arXiv preprint 2107.06126, 2021.

[8] Chetupalli, S.R., et al., Multi-modal Point-of-Care Diagnostics for COVID-19 Based On Acoustics and Symptoms. arXiv preprint 2106.00639, 2021.

[9] Muguli, A., et al., DiCOVA Challenge: Dataset, task, and baseline system for COVID-19 diagnosis using acoustics. arXiv preprint 2103.09148, 2021.

[10] Jayachitra, V.P., et al., A cognitive IoT-based framework for effective diagnosis of COVID-19 using multimodal data. Biomed Signal Process Control, 2021. 70: p. 102960.

[11] Pahar, M., et al., COVID-19 cough classification using machine learning and global smartphone recordings. Comput Biol Med, 2021. 135: p. 104572.

[12] Tena, A., F. Claria, and F. Solsona, Automated detection of COVID-19 cough. Biomed Signal Process Control, 2022. 71: p. 103175.

[13] Amoh, J. and K. Odame, Deep Neural Networks for Identifying Cough Sounds. IEEE Trans Biomed Circuits Syst, 2016. 10(5): p. 1003–1011.

[14] Bartl-Pokorny, K.D., et al., The voice of COVID-19: Acoustic correlates of infection in sustained vowels. The Journal of the Acoustical Society of America, 2021. 149(6): p. 4377–4383.

[15] Hoyos-Barcelo, C., et al., Efficient k-NN Implementation for Real-Time Detection of Cough Events in Smartphones. IEEE J Biomed Health Inform, 2018. 22(5): p. 1662–1671.

[16] Knocikova, J., et al., Wavelet analysis of voluntary cough sound in patients with respiratory diseases. J Physiol Pharmacol, 2008. 59(Suppl 6): p. 331–40.

[17] Liu, J.-M., et al. Cough detection using deep neural networks. in 2014 IEEE International Conference on Bioinformatics and Biomedicine (BIBM). 2014. IEEE.

[18] Monge-Alvarez, J., et al., Robust Detection of Audio-Cough Events Using Local Hu Moments. IEEE J Biomed Health Inform, 2019. 23(1): p. 184–196.

[19] Monge-Alvarez, J., et al., Effect of importance sampling on robust segmentation of audio-cough events in noisy environments. Annu Int Conf IEEE Eng Med Biol Soc, 2016. 2016: p. 3740–3744.

[20] Monge-Álvarez, J., et al., A machine hearing system for robust cough detection based on a high-level representation of band-specific audio features. IEEE Transactions on Biomedical Engineering, 2018. 66(8): p. 2319–2330.

[21] Pal, A. and M. Sankarasubbu. Pay attention to the cough: Early diagnosis of COVID-19 using interpretable symptoms embeddings with cough sound signal processing. in Proceedings of the 36th Annual ACM Symposium on Applied Computing. 2021.

[22] Pinkas, G., et al., SARS-CoV-2 detection from voice. IEEE Open Journal of Engineering in Medicine and Biology, 2020. 1: p. 268–274.

[23] Belkacem, A.N., et al., End-to-End AI-Based Point-of-Care Diagnosis System for Classifying Respiratory Illnesses and Early Detection of COVID-19: A Theoretical Framework. Front Med (Lausanne), 2021. 8: p. 585578.

[24] Coppock, H., et al., End-to-end convolutional neural network enables COVID-19 detection from breath and cough audio: a pilot study. BMJ Innov, 2021. 7(2): p. 356–362.

[25] Lella, K.K. and A. Pja, Automatic COVID-19 disease diagnosis using 1D convolutional neural network and augmentation with human respiratory sound based on parameters: cough, breath, and voice. AIMS Public Health, 2021. 8(2): p. 240–264.

[26] Shimon, C., et al., Artificial intelligence enabled preliminary diagnosis for COVID-19 from voice cues and questionnaires. J Acoust Soc Am, 2021. 149(2): p. 1120.

[27] Mouawad, P., T. Dubnov, and S. Dubnov, Robust Detection of COVID-19 in Cough Sounds. SN Computer Science, 2021. 2(1): p. 1–13.

[28] Orlandic, L., T. Teijeiro, and D. Atienza, The COUGHVID crowdsourcing dataset, a corpus for the study of large-scale cough analysis algorithms. Sci Data, 2021. 8(1): p. 156.

[29] Bhateja, V., A. Taquee, and D.K. Sharma. Pre-processing and classification of cough sounds in noisy environment using SVM. in 2019 4th International Conference on Information Systems and Computer Networks (ISCON). 2019. IEEE.

[30] Laguarta, J., F. Hueto, and B. Subirana, COVID-19 artificial intelligence diagnosis using only cough recordings. IEEE Open Journal of Engineering in Medicine and Biology, 2020. 1: p. 275–281.

[31] Imran, A., et al., AI4COVID-19: AI enabled preliminary diagnosis for COVID-19 from cough samples via an app. Inform Med Unlocked, 2020. 20: p. 100378.

[32] Cohen-McFarlane, M., R. Goubran, and F. Knoefel. Comparison of silence removal methods for the identification of audio cough events. in 2019 41st Annual International Conference of the IEEE Engineering in Medicine and Biology Society (EMBC). 2019. IEEE.

[33] Cohen-McFarlane, M., R. Goubran, and F. Knoefel, Novel coronavirus cough database: NoCoCoDa. IEEE Access, 2020. 8: p. 154087–154094.

[34] Miranda, I.D., A.H. Diacon, and T.R. Niesler. A comparative study of features for acoustic cough detection using deep architectures. in 2019 41st Annual International Conference of the IEEE Engineering in Medicine and Biology Society (EMBC). 2019. IEEE.

[35] Mydukuri, R.V., et al., Deming least square regressed feature selection and Gaussian neuro-fuzzy multi-layered data classifier for early COVID prediction. Expert Syst, 2021: p. e12694.

[36] Mohammed, E.A., et al., An ensemble learning approach to digital corona virus preliminary screening from cough sounds. Sci Rep, 2021. 11(1): p. 15404.

[37] Feng, K., et al. Deep-learning Based Approach to Identify Covid-19. in SoutheastCon 2021. 2021. IEEE.

[38] Melek, M., Diagnosis of COVID-19 and non-COVID-19 patients by classifying only a single cough sound. Neural Comput Appl, 2021: p. 1–12.

[39] Melek Manshouri, N., Identifying COVID-19 by using spectral analysis of cough recordings: a distinctive classification study. Cognitive Neurodynamics, 2021.

[40] Quatieri, T.F., T. Talkar, and J.S. Palmer, A framework for biomarkers of COVID-19 based on coordination of speech-production subsystems. IEEE Open Journal of Engineering in Medicine and Biology, 2020. 1: p. 203–206.

[41] Chowdhury, M.E., et al., QUCoughScope: An Artificially Intelligent Mobile Application to Detect Asymptomatic COVID-19 Patients using Cough and Breathing Sounds. arXiv preprint 2103.12063, 2021.

[42] Pahar, M., et al., COVID-19 Detection in Cough, Breath and Speech using Deep Transfer Learning and Bottleneck Features. arXiv preprint 2104.02477, 2021.

[43] Sharma, M., et al., Impact of data-splits on generalization: Identifying COVID-19 from cough and context. arXiv preprint 2106.03851, 2021.

[44] Sharma, N., et al., Coswara--A Database of Breathing, Cough, and Voice Sounds for COVID-19 Diagnosis. arXiv preprint 2005.10548, 2020.

[45] Xue, H., & Salim, F. D., Exploring Self-Supervised Representation Ensembles for COVID-19 Cough Classification. arXiv preprint 2105.07566., 2021.

[46] Akman, A., et al., Evaluating the COVID-19 Identification ResNet (CIdeR) on the INTERSPEECH COVID-19 from Audio Challenges. arXiv preprint 2107.14549, 2021.

[47] Bagad, P., et al., Cough against COVID: Evidence of COVID-19 signature in cough sounds. arXiv preprint 2009.08790, 2020.

[48] Brown, C., et al., Exploring automatic diagnosis of COVID-19 from crowdsourced respiratory sound data. arXiv preprint 2006.05919, 2020.

[49] Shams, A.B., et al., Telehealthcare and Covid-19: A Noninvasive & Low Cost Invasive, Scalable and Multimodal Real-Time Smartphone Application for Early Diagnosis of SARS-CoV-2 Infection. arXiv preprint 2109.07846, 2021.

[50] Nguyen, L.H., et al., Fruit-CoV: An Efficient Vision-based Framework for Speedy Detection and Diagnosis of SARS-CoV-2 Infections Through Recorded Cough Sounds. arXiv preprint 2109.03219, 2021.

[51] Chowdhury, N.K., M.A. Kabir, and M. Rahman, An Ensemble-based Multi-Criteria Decision Making Method for COVID-19 Cough Classification. arXiv preprint 2110.00508, 2021.

[52] Schuller, B.W., et al., The INTERSPEECH 2021 Computational Paralinguistics Challenge: COVID-19 cough, COVID-19 speech, escalation & primates. arXiv preprint 2102.13468, 2021.

[53] Schuller, B.W., H. Coppock, and A. Gaskell, Detecting COVID-19 from breathing and coughing sounds using deep neural networks. arXiv preprint 2012.14553, 2020.

[54] Bales, C., et al. Can machine learning be used to recognize and diagnose coughs? In 2020 International Conference on e-Health and Bioengineering (EHB). 2020. IEEE.

[55] Chen, X., M. Hu, and G. Zhai, Cough Detection Using Selected Informative Features from Audio Signals. arXiv preprint 2108.03538, 2021.

[56] Deshpande, G. and B.W. Schuller, Audio, Speech, Language, & Signal Processing for COVID-19: A Comprehensive Overview. arXiv preprint 2011.14445, 2020.

[57] Pizzo, D.T., S. Esteban, and M. Scetta, IATos: AI-powered pre-screening tool for COVID-19 from cough audio samples. arXiv preprint 2104.13247, 2021.

[58] Hossain, M.Z., M.B. Uddin, and K.A. Ahmed, CovidEnvelope: A Fast Automated Approach to Diagnose COVID-19 from Cough Signals. medRxiv, 2021.

[59] Ritwik, K.V.S., S.B. Kalluri, and D. Vijayasenan, COVID-19 patient detection from telephone quality speech data. arXiv preprint 2011.04299, 2020.

[60] Coppock, H., et al., COVID-19 detection from audio: seven grains of salt. Lancet Digit Health, 2021. 3(9): p. e537–e538.

[61] Khanzada, A., et al., Challenges and Opportunities in Deploying COVID-19 Cough AI Systems. J Voice, 2021.

[62] Adans-Dester, C.P., et al., Can mHealth technology help mitigate the effects of the COVID-19 pandemic? IEEE Open Journal of Engineering in Medicine and Biology, 2020. 1: p. 243–248.

[63] Faezipour, M. and A. Abuzneid, Smartphone-based self-testing of COVID-19 using breathing sounds. Telemedicine and e-Health, 2020. 26(10): p. 1202–1205.

[64] Schuller, B.W., et al., COVID-19 and Computer Audition: An Overview on What Speech & SoundAnalysis Could Contribute in theSARS-CoV-2 Corona Crisis. Frontiers in Digital Health, 2021. 3: p. 14.

[65] Ting, D.S.W., et al., Digital technology and COVID-19. Nature medicine, 2020. 26(4): p. 459–461.

[66] Topol, E.J., Is my cough COVID-19? Lancet, 2020. 396(10266): p. 1874.

